# Adherence to Anti-Epileptic Drugs and Self-reported Availability and Affordability of the Drugs in Addis Ababa, Ethiopia

**DOI:** 10.1101/2024.02.21.24303153

**Authors:** Bethlehem Shawel, Yemane Berhane

**Author notes:** Corresponding author: (BS).

## Abstract

**Background:** Anti-epileptic drugs (AEDs) are the primary therapeutic mode to control seizures in patients with epilepsy. Adherence to the medications is critical to achieving the goals of epilepsy therapy. However, the cost of the drugs and interrupted availability of AEDs contribute to non-adherence to epilepsy treatment. Therefore, this study aimed to assess AED adherence and its association with self-reported drug availability and affordability.

**Objective:** To assess whether self-reported availability and affordability of Antiepileptic drugs affect drug adherence among Epileptic Patients at Eka Kotebe General Hospital, Addis Ababa, Ethiopia, from January 2023 to March 2023.

**Methods:** A hospital-based analytical cross-sectional study was conducted among 357 epileptic patients using the Consecutive sampling method in Eka Kotebe General Hospital, Addis Ababa, Ethiopia. AED adherence was measured using a self-report 3items questionnaire focusing on medication use patterns of patients from their last visit to the current visit. Statistical packages for Social Sciences 26.0 version statistical software cleaned, coded and analyzed the collected data. Binary logistic regression was fitted, and P-values less than 0.05 were considered to have statistical significance.

**Result:** The prevalence of AED adherence was 55.2% with 95% CI (50.1%; 60.2%). About two-thirds (61.3%) of patients in this study had limited access to the AEDs or could not afford the medications. Self-reported availability of AEDs (AOR=2.04, 95% CI=1.03, 4.03) was significantly associated with AED adherence. Self-reported affordability of AEDs was associated with AED adherence (COR=1.59, CI=1.04, 2.42, P-Value=0.031) in the Bivariate logistic regression analysis; however, when adjusted for other covariates in the multivariable logistic regression, no significant association was observed (p=0.730).

**Conclusion and Recommendation:** Only about half of the epileptic patients adhered to AEDs at Eka Kotebe General Hospital. Self-reported availability of AEDs was an essential factor. Improving access to AEDs is critical to improving adherence and management of epilepsy.

## Introduction

Medication adherence is the extent to which individuals take their medications as prescribed with respect to dosage and dosage intervals (1). In both developed and developing countries, non-adherence to medication remains a significant concern for healthcare providers as well as patients because of its adverse consequences on therapeutic outcomes. It is significant, especially in chronic illnesses involving complex and long-term medical regimens (2).

Epilepsy is a common chronic neurological disease of the brain affecting 50 million people worldwide, of whom 40 million are estimated to live in developing countries (3). Antiepileptic Drugs (AEDs) are the mainstay of long-term epilepsy treatment, and they are 70% effective at reducing epileptic attacks in adults (4). However, about 90% of epileptic patients in developing countries are not receiving appropriate treatment due to cultural attitudes, lack of prioritization, poor healthcare system, and inadequate supply of antiepileptic drugs (AEDs) (5).

In Ethiopia, epilepsy is often mistakenly seen as a form of mental illness by the Community and is usually treated by psychiatrists and psychiatric nurses. The most prescribed Antiepileptic drugs are phenobarbitone, phenytoin, and sometimes carbamazepine and sodium Valproate.

A study in Western countries revealed an adherence rate of 71% in the US (6) and 59% in the UK (7). The result was not different in Gondar, Ethiopia (70.8%) (8) and Jimma, Ethiopia (58.5%) (9). On the other hand, the report was higher in India (98.6%) (10) and Palestine (85.3%) (11). However, a lower adherence rate was found in China 51.9% (12), Nigeria (32.6%) (13), and South Africa (54.6%) (14).

The consequence of AED nonadherence behavior has been associated with poor seizure control, increased morbidity and mortality along with increased time of hospitalization, worsened patient outcome, poor quality of life, and increased healthcare cost. AED nonadherence will also lead to an increase in the burden of inpatient and emergency department services; moreover, it also affects the family members socially, economically, and psychologically (15).

Availability and affordability of AEDs are poor and act as a barrier to accessing treatment for epilepsy in low and middle-income countries (16). In most low-income countries, access to medicines remains very low. The availability and affordability of drugs are two key factors that affect patients’ access to treatment. A study of the availability and prices of AEDs in southern Vietnam showed that only 57% of the public and private pharmacies surveyed had AEDs available (17). A second study conducted in Zambia found that nearly one-half of the government, private, and nongovernmental organization (NGO) pharmacies surveyed did not carry AEDs (18). There is a failure to explore these factors, particularly in Ethiopia.

A study done in Bangui, Central African Republic, on the Problem of accessibility to anti-seizure drugs for patients who have epilepsy, shows that the lack of trained personnel, the inadequacy of pharmaceutical structures, the insufficient availability of antiepileptic drugs, and their very high cost are factors limiting the accessibility of antiepileptic medications for epileptic patients (19).

This study aimed to evaluate drug availability and affordability from the patient’s point of view. This allows for a more direct assessment of the role of these factors in adherence. However, the studies and tools developed thus far were from a system perspective. Hence, this study aims to assess the association of self-reported drug availability and affordability with drug adherence. In addition, AED adherence was also assessed to fill the gaps in epilepsy treatment.

## Methodology

### Study design and period

A hospital-based Analytical cross-sectional study with internal comparison was conducted from January 2023 to March 2023.

### Study Setting

The study was conducted at Eka Kotebe General Hospital, located in the eastern part of Addis Ababa, the capital city of Ethiopia. Eka Kotebe General Hospital was established as an extension of Amanuel Mental Specialized Hospital until April 2020, when it became a stand-alone federal hospital. The hospital has a dedicated clinic for patients with epilepsy. There were about 2478 epileptic patients who had regular follow-ups in a year at the outpatient department, and on average, 200 epilepsy patients have follow-ups at the hospital. Patients pay for the services they get from the hospital and for purchasing AEDs, except those who use CBHI (community-based health insurance). The hospital also gives psychiatric services at the outpatient level. In addition, the hospital has served as a dedicated inpatient COVID treatment center.

### Source population

Adult epileptic patients who have been on treatment with one or more antiepileptic drugs and who had follow-ups at the outpatient units during the study period.

### Study population

We selected adult epileptic patients who have been on treatment with one or more Antiepileptic drugs and who had follow-up at the outpatient units during the study period, fulfilling the inclusion criteria.

### Inclusion criteria

Adult Epileptic Patients (age≥18 years) who had a follow-up in the hospital for at least three months and received AEDs at the last visit were eligible for the study. Patients with general severe medical conditions and unable to communicate were excluded.

### Sample Size and Sampling Technique

The sample size for the first objective was determined using a single population proportion formula considering the following assumptions (z=1.96, d= 0.05, and P=65%) (20). Second, an attempt was made to calculate the sample size by considering the association of Drug availability and affordability with adherence using Open Epi version 2.3.1. Finally, the sample size we obtained from the first objective was the largest and was taken as our final sample. By considering a non-response rate of 10%, the final estimated sample size was 385.

Consecutive sampling was used to recruit samples for the study each day of the data collection process until the desired sample size was obtained.

### Data collection tool and procedures

Data was collected using a structured questionnaire translated into a local language (Amharic) and pre-tested. Trained nurses interviewed volunteer participants face-to-face during their follow-up clinic visits. The data was collected after explaining the purpose of the study to the participants and earning informed written consent. The patient interview aimed to gather information related to sociodemographic, clinical factors, psychosocial factors, self-reported drug availability, affordability, and adherence. Their medical records were reviewed for the type of seizure and prescribed AEDs.

**Adherence** of epileptic patients to their AEDs was assessed using a self-reported questionnaire; in this study, a 3 items questionnaire focusing on medication use patterns of patients from their last visit to the current visit was used. Patients were asked 3questions: “Did you forget to take your medicine since your last visit?” “In the past two weeks, were there any days you did not take your medicine?” and “Did you take all your medicine yesterday?” The response choices were yes/no, and a response “no” was rated as “1,” and a “yes” response as “0,” except for item 3. For item 3, a response “yes” was rated as “1” and “no” as “0”. The total score ranges from 0 to 3, with scores of </=1, 1 to <3, and 3 reflecting low, medium, and high adherence, respectively. For data analysis, scores were categorized into two, scores <3 were assigned as non-adherent, and those who scored 3 were assigned as adherent.

### Data quality control

To ensure the data quality, a data collection tool was prepared after reviewing the literature related to the study. Data was collected by 4 BSC nurses who know about epilepsy and a supervisor. A thorough training was given by the principal investigator about the general objective of the study and the contents of the questionnaire. The questionnaire was pre-tested a week before the data collection started on 10 % of the sample size at Eka Kotebe General Hospital Outpatient units. After a thorough and deep review of inputs obtained during the pre-test, the final tool was developed with some modifications. The pre-tested patients were excluded from the analysis. Data collectors were supervised daily, and the supervisor and principal investigator checked the filled questionnaires daily.

### Data analysis

The gathered quantitative data was cleaned, arranged, coded, and then analyzed through statistical packages for social sciences (SPSS) version 26.0 statistical Software. Categorical variables were expressed by percentage and frequency, whereas numerical variables were present by median with Inter Quartile range (IQR).

The association between the independent variables and Adherence to AEDs was analyzed using a binary logistic regression model. Bivariate binary logistic regression was run at a 25% significance level to screen out potentially significant independent variables. A multivariable binary logistic regression model was run by including the significant independent variables from the bivariate binary logistic regression model. To measure the presence and strength of association between the independent variables and adherence to AEDs, adjusted odds ratio (AOR), P-value, and 95% CI for AOR were calculated separately for self-reported availability and affordability of AEDs. Since the ORs were not different, in the final model, the two variables were considered together, and variables with a P-value of ≤ 0.05 were considered significantly associated with adherence to AEDs.

### Operational definitions

#### Adherence

In this study, AED adherence was measured using a self-report 3-item questionnaire focusing on medication use patterns of patients from their last visit to the current visit, which was developed by modifying the Morisky Medication Adherence Scale-8 (MMAS-8).

#### Self-reported Drug Availability

In this study, Drug availability was measured from the patient’s perspective based on their ability to obtain their medication in the same facility where treatment was given, a nearby facility, or near their residence. Accordingly, Drug Availability was assigned as “**Available**” if patients were able to gain drugs in the same or nearby facility where they gain treatment or near their residency and “**Unavailable**” if they were not able to gain drugs neither in the same or nearby pharmacy/ facility where treatment was given nor nearby pharmacy/ facility to their residence.

#### Self-reported Drug Affordability

In this study, Drug affordability was assigned as “**Affordable**” if patients could pay for drugs with no difficulty or gain medications free and as “**Unaffordable**” if patients were paying for medications with difficulty.

#### AED (Antiepileptic Drugs)

AEDs are the primary medications used to control seizures in epileptic patients. In our setup, the most prescribed antiepileptic drugs are phenobarbitone, phenytoin, and sometimes carbamazepine and sodium valproate. In this study, the number of AEDs used by patients were classified as monotherapy and ploy-therapy, implying patients who are using single and double or multiple antiepileptic drugs, respectively.

### Ethical consideration

The research and ethical review board of the Addis Continental Institute of Public Health granted ethical clearance and approval of the study. Following the approval, Eka Kotebe General Hospital Institutional Review Board (IRB) obtained official permission. Before data collection, informed written consent was obtained from the study participants. Individuals were told that they had a right to withdraw from the study at any time, and this would not affect the service they got from the hospital. Confidentiality was ensured during the patient interviews and the review of charts.

## Results

### Socio-Demographic characteristics

A total of 357 participants participated in the study, with a response rate of 92.7%. From this, 160(44.8%) of the respondents were 15-29 years old. More than half of the study participants (61.1%) were single and 184 (51.5%) were males. Regarding educational status, the majority of the study participants had primary and secondary school, 95 (26.6%) and 109(30.5%), respectively. Around two-thirds of the study participants were unemployed and, 191 (53.5%) were living in Addis Ababa, and nearly a third were within the same sub-city. The median (IQR) family size of the participants was 4 (3, 5) (Table 1).

**Table 1:** Socio-Demographic Characteristics of Adult Epileptic Patients in Eka Kotebe General Hospital, Ethiopia, 2023 (n=357)

| Variable | Category | Frequency | Percentage |
| --- | --- | --- | --- |
| **Family size (Median, IQR)** |  |  | 4 (3,5) |
| **Age(Year)** | 15-29 | 160 | 44.8 |
|  | 30-44 | 123 | 34.5 |
|  | >45 | 74 | 20.7 |
| **Sex** | Female | 173 | 48.5 |
|  | Male | 184 | 51.5 |
| **Marital Status** | Single | 218 | 61.1 |
|  | Married | 139 | 38.9 |
| **Educational Status** | No formal education | 80 | 22.4 |
|  | Primary education (1-8) | 95 | 26.6 |
|  | Secondary education (9-12) | 109 | 30.5 |
|  | College and above | 73 | 20.4 |
| **Usual place of residence** | Within the same sub-city | 109 | 30.5 |
|  | Addis Ababa | 191 | 53.5 |
|  | Outside Addis Ababa | 57 | 16 |
| **Employment Status** | Gainfully Employed | 130 | 36.4 |
|  | Unemployed | 227 | 63.6 |
| **Social Support** | Not often | 87 | 24.4 |
|  | Sometimes | 152 | 42.6 |
|  | Often | 118 | 33.1 |
| **Financial support** | No | 181 | 71.5 |
|  | Yes | 72 | 28.5 |

The majority of the respondents, 181(71.5%), had no financial support, and 118 (33.1%) thought they had someone around to help them often when they needed help. (Table 1).

### Clinical factors for antiepileptic drug adherence

Among the study participants, the majority of the patients, 279 (78.2%), were diagnosed with a Generalized type of seizure, and more than half, 227(63.6%) of the participants were on monotherapy. The median (IQR) age for the onset of the illness of the participants was 17(10, 24) years, and about slightly two-thirds, 223(62.5%), of the participants had a duration of illness of greater than/equal to 11 years. While co-morbidity was found in less than a fifth of the participants and a small number, 17(4.8%) of the participants had an adverse effect that prompted them to stop taking medication since their last visit (Table 2).

**Table 2:** Clinical factors for antiepileptic drug adherence among adult Epileptic patients at Eka Kotebe General Hospital, Ethiopia, 2023 (n=357)

| Variable | Category | Frequency | Percentage |
| --- | --- | --- | --- |
| **Age of onset of Epilepsy (Year) (Median, IQR)** |  |  | 17(10,24) |
| **Duration of illness (in Years)** | <5 years | 48 | 13.4 |
|  | 6-10 years | 82 | 23 |
|  | >11 years | 223 | 62.5 |
| **Types of Seizure** | Generalized | 279 | 78.2 |
|  | Partial | 66 | 18.5 |
|  | Unclassified | 12 | 3.4 |
| **Number of AEDs** | Monotherapy | 227 | 63.6 |
|  | Polytherapy | 130 | 36.4 |
| **History of Adverse effects of AEDs that prompt to stop taking medication** | No | 283 | 79.3 |
|  | Yes | 74 | 20.7 |
| **History of Adverse effects of AEDs that prompt to stop taking medication since the last visit** | No | 340 | 95.2 |
|  | Yes | 17 | 4.8 |
| **Comorbidity** | No | 308 | 86.3 |
|  | Yes | 49 | 13.7 |

### Prevalence of adherence to AEDs and Self-reported drug availability and affordability among adult Epileptic patients

The prevalence of AED adherence was 55.2% with 95% CI (50.1%; 60.2%). In this study, self-reported availability and affordability of AEDs were found to be 80.1% and 53.2% respectively (Table 3).

**Table 3:** Prevalence of adherence to AEDs and Self-reported drug availability and affordability among adult Epileptic patients at Eka Kotebe General Hospital, Ethiopia, 2023 (n=357)

| **Variable** | **Category** | **Frequency** | **Percentage** |
| --- | --- | --- | --- |
| **Adherence to AEDs** | Non-Adherent | 160 | 44.8 |
|  | Adherent | 197 | 55.2 |
| **Self-reported Availability of AEDs** | Unavailable | 71 | 19.9 |
|  | Available | 286 | 80.1 |
| **Self-reported Affordability of AEDs** | Unaffordable | 167 | 46.8 |
|  | Affordable | 190 | 53.2 |

The unavailability of the medications at hand 36.3% and forgetfulness to take medicine 35.6%, were the most common reasons for medication nonadherence, followed by drug unaffordability 25%, and travel outside the home 13.1% was the least reasons for medication non-adherence (Fig 1).

**Fig 1.**
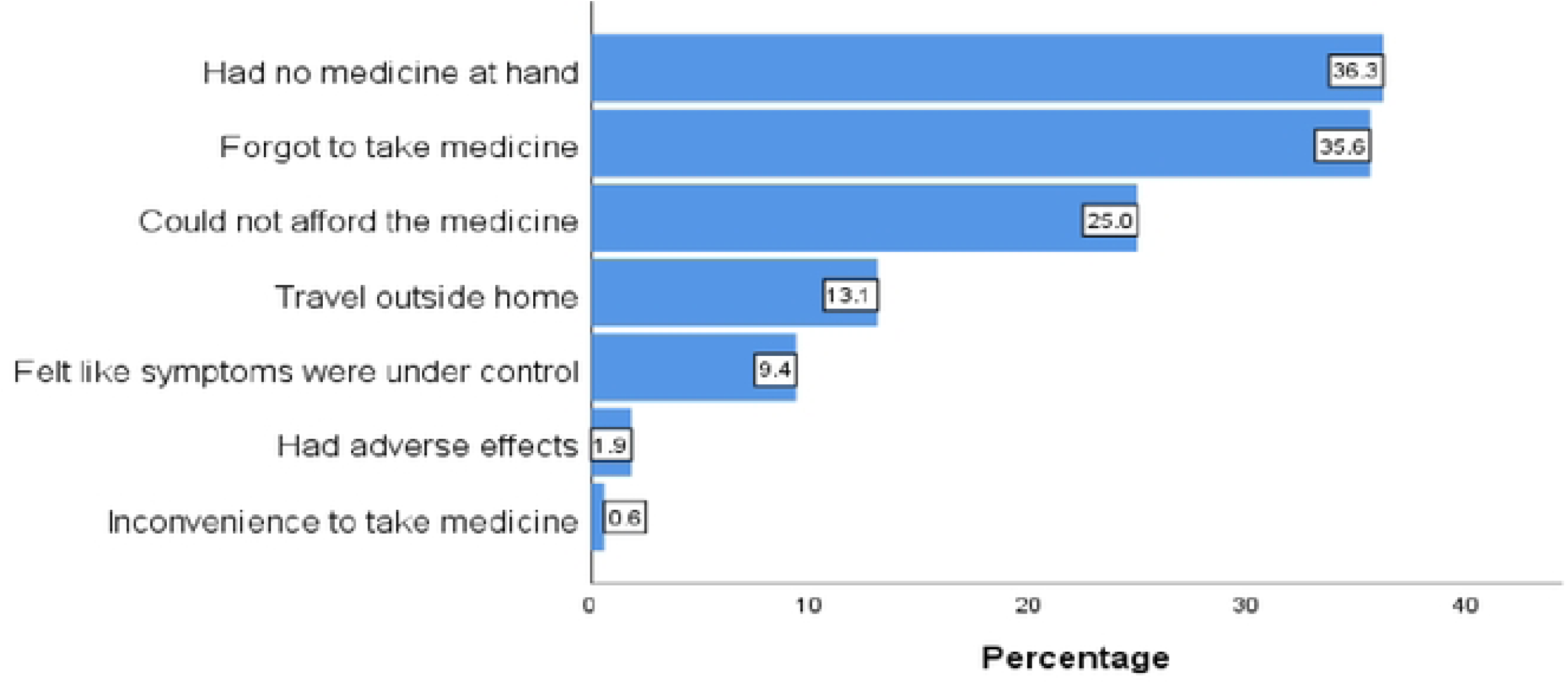
Reasons for AEDs non-adherence at Eka Kotebe General Hospital, Ethiopia, 2023 (n= 357)

### Association of Self-reported drug availability and affordability with anti-epileptic drug adherence among people with epilepsy

In the Bivariate binary logistic regression, Educational status, Number of AEDs, and self-reported affordability of AEDs were significantly associated with adherence to AEDs. However, on the multivariable binary logistic regression model, after adjusting for the covariates, Patients who finished secondary school, were educated up to college level and above, Number of AEDs, and self-reported Availability of AEDs were found to have a statistically significant relationship with adherence to AEDs at 5% level of significance.

Accordingly, after adjusting for other covariates, the odds of being adherent to AEDs among those with secondary education were 2.2times higher as compared to those with no formal education **(AOR=2.24, 95% CI=1.08, 4.61, p-value=0.030).** Similarly, the odds of adherent to AEDs were 2.7 times higher for those educated up to college level and above than those with no formal education **(AOR= 2.71, 95% CI= 1.14, 6.46, p-value=0.025)**. Regarding self-reported availability of AEDs, the odds of being adherent to AEDs were 2times higher among those who reported the medications available as compared with those who reported unavailable **(AOR=2.04,95% CI=1.03,4.03, p-value=0.041).** On the other hand, the odds of being adherent to AEDs were 45.5% less for those on polytherapy than those on monotherapy **(AOR= 0.55, 95% CI= 0.31,0.95, p-value=0.031).** In bivariate logistic regression analysis, self-reported affordability of AEDs was associated with AEDs adherence (COR=1.59, CI=1.04, 2.42, P-Value=0.031); however, when adjusted for other covariates in the multivariable logistic regression, no significant association was observed (p=0.730) (Table 4).

**Table 4:** Bivariate and Multivariate analysis of variables associated with AED adherence (n=357)

| Variables | Category | Adherence to AEDs |  | COR (95% CI) | AOR (95% CI) | P-Value |
| --- | --- | --- | --- | --- | --- | --- |
|  |  | Non-adherent (n) | Adherent (n) |  |  |  |
| Age (years) | 15-29 | 72 | 88 | 1 | 1 |  |
|  | 30-44 | 55 | 68 | 1.01(0.63-1.62) | 1.08(0.59-1.97) | 0.808 |
|  | >45 | 33 | 41 | 1.02(0.59-1.77) | 0.95(0.45-2.00) | 0.884 |
| Sex | Female | 77 | 96 | 1 | 1 |  |
|  | Male | 83 | 101 | 0.98(0.64-1.48) | 0.89(0.53-1.50) | 0.658 |
| Educational Status | No formal education | 43 | 37 | 1 | 1 |  |
|  | Primary education (1-8) | 50 | 45 | 1.05(0.58-1.90) | 1.12(0.54-2.33) | 0.767 |
|  | Secondary education (9-12) | 41 | 68 | **1.93(1.07-3.46)** | **2.24(1.08-4.61)** | **0.030\*** |
|  | College and above | 26 | 47 | **2.10(1.10-4.03)** | **2.71(1.14-6.46)** | **0.025\*** |
| **Number of Antiepileptic drugs** | Monotherapy | 89 | 138 | 1 | 1 |  |
|  | Polytherapy | 71 | 59 | **0.54(0.35-0.83)** | **0.55(0.31-0.95)** | **0.031\*** |
| **Comorbidity** | No | 137 | 171 | 1 | 1 |  |
|  | Yes | 23 | 26 | 0.91(0.50-1.66) | 0.79(0.34-1.82) | 0.579 |
| **Self-reported Availability of AEDs** | Unavailable | 36 | 35 | 1 | 1 |  |
|  | Available | 124 | 162 | **1.34 (0.80-2.26)** | **2.04(1.03-4.03)** | **0.041\*** |
| **Self-reported Affordability of AEDs** | Unaffordable | 85 | 82 | 1 | 1 |  |
|  | Affordable | 75 | 115 | 1.59(1.04-2.42) | 1.11(0.63-1.96) | 0.730 |
| **Financial Support** | No | 89 | 92 | 1 | 1 |  |
|  | Yes | 34 | 38 | 1.08(0.63-1.87) | 0.93(0.52-1.66) | 0.799 |

\* Statistically Significant at P<0.05

## Discussion

Findings of adherence in this study showed that less than two-thirds (55.2%; 95% CI (50.1%-60.2%) of the study participants were adherent to antiepileptic drugs. Globally, the adherence rate ranges between 33 and 99 % (6,8,10,12,13).

The prevalence of adherence to antiepileptic drugs in this study was in line with a cross-sectional study conducted in Addis Ababa (52.4%) (21), Jimma (58.5 %) (9), and Nigeria (55.5 %) (22). However, it was lower than the study done in Gondar, Ethiopia (61.5%)(23), Southwestern Ethiopia (63.5%)(24), and India (72.3%) (25). The probable reason for these discrepancies could be the differences in the methodology used for assessing adherence rate and the study design. For example, a prospective observational study was employed in the study from southwestern Ethiopia, and adherence was assessed by the Hill–Bone compliance to the high blood pressure therapy scale (24), but the current study employed a cross-sectional study design, and adherence was assessed using self-report questionnaire modified from MMAS 8. The other reason could be due to the socio-demographic characteristics of the study participants as well as the study area.

On the other hand, the findings of this study were higher than the studies done in Brazil and Nigeria, which were (33.8%) (26) and (31.6%) (13), respectively, and this difference was probably due to the difference in AEDs multidrug treatment. For instance, 71.1% of people with epilepsy in Brazil and 85% in Nigeria were on multiple AED treatment, while in the current study, only 36.4% of people with epilepsy were in poly-AED treatment.

The most common reasons for missing doses in this study were the unavailability of the medications at hand (36.3%) followed by forgetfulness to take medicine (35.6%), which is supported by the findings of studies in Amanuel Mental Specialized Hospital, Ethiopia (27), Jimma Ethiopia (24) and Nigeria (22).

In this study, factors found to have a significant association with adherence to AEDs were higher educational status, self-reported availability of AEDs, and being on polytherapy. This study revealed the odds of being adherent to AEDs were 2.7 times higher for those who were educated up to college level and above than those with no formal education. This result aligns with the study conducted in India (28). This might be due to patients who are more educated might question their health care providers about their disease and its medications, might be more aware of the disease progression and follow the prescription and instructions as told Whereas, those who are less educated in our setting might prefer other alternative traditional practices at home than proper medication follow up.

Besides, Patients taking combination drugs were less likely to Adhere. Polytherapy increases the potential for drug-drug interactions, results in failure to evaluate the individual drugs may affect compliance and is associated with a higher cost of medication and the necessity for therapeutic drug monitoring (29). Our study revealed the odds of being adherent to AEDs were 45.5% less for those on polytherapy as compared with those who were on monotherapy. Similar studies found a negative association between the complexity of medication and patient adherence (22,28). Regarding self-reported availability of AEDs, the odds of being adherent to AEDs were two times higher among those who reported the medications available as compared with those who reported unavailable. This finding was supported by the study from Jimma, Ethiopia (24), and India (28). In bivariate logistic regression analysis, self-reported affordability of AEDs was associated with AED adherence. This finding was supported by the study from South Western Ethiopia (30). The possible justification was that patients who got their medication out of pocket would sometimes cease to purchase their medications, and those who got free will continue their medications in whatever circumstances.

By contrast, the patient’s adherence to their medication did not show a significant association with age and gender. This result is in line with the findings in Gondar, Ethiopia (23), Nigeria (22), and India (28).

This study has a few limitations. First, it was a cross-sectional study with no prospective follow-up period. As a result, it may be subjected to recall bias. Second, a self-reported measure of adherence, which is prone to social desirability bias, was used to measure adherence. Third, it was conducted in a single setting, which makes it difficult to generalize. Therefore, a large-scale nationwide study is highly recommended.

## Conclusion and Recommendation

In conclusion, overall, only about half of the patients adhered to Antiepileptic drugs at Eka Kotebe General Hospital. The self-reported availability of AEDs was significantly associated with adherence to antiepileptic drugs. Improving access to AEDs is critical to improving adherence and management of epilepsy.

## Data Availability

All relevant data are within the manuscript and its Supporting Information files.

## Acknowledgments

We thank the patients for their consent to participate in this study and their patience during the interview. We also appreciate the staff of Eka Kotebe General Hospital for their cooperation and support throughout the data collection process.

## Abbreviations

AEDs: Anti-Epileptic Drugs
AOR: Adjusted Odds Ratio
CBHI: Community-Based Health Insurance CI Confidence Interval
COR: Crudes Odds Ratio
IQR: Inter Quartile Range
IRB: Institutional Review Board
MMAS-8: Morisky Medication Adherence Scale-8 OR Odds ratio
SPSS: Statistical Package for Social Sciences

## Supporting Information

**S1 File. English Version of Questionnaires**

**S2 File. Amharic Version of Questionnaires**

